# Modeling of leptospirosis outbreaks in relation to hydroclimatic variables in the northeast of Argentina

**DOI:** 10.1101/2021.07.06.21260095

**Authors:** Andrea A. Gómez, María Soledad López, Gabriela V. Müller, Leonardo López, Walter Sione, Leonardo Giovanini

## Abstract

The transmission of leptospirosis is conditioned by climatic variables. In northeastern Argentina leptospirosis outbreaks occur mainly in coincidence with periods of abundant precipitation and high hydrometric level. A Susceptible-Infectious-Recovered Epidemiological Model (SIR) is proposed, which incorporates hydroclimatic variables for the three most populated cities in the area (Santa Fe, Paraná and Rosario), during the 2009 – 2018 period. Results obtained by solving the proposed SIR model for the 2010 outbreaks are in good agreement with the actual data, capturing the dynamics of the leptospirosis outbreak wave. However, the model does not perform very well when isolated cases appear outside the outbreak periods, probably due to non-climatic factors not explicitly considered in the present version of the model. Nevertheless, the dynamic modeling of infectious diseases considering hydroclimatic variables constitutes a climatic service for the public health system, not yet available in Argentina.

## 1. Introduction

Leptospirosis is a zoonosis found worldwide and it is a major public health issue in many rural and urban surroundings in temperate and tropical climates. This vector disease is present especially in many countries of Latin America and South-East Asia. No vaccine is available, so prevention is largely dependent on sanitation measures that may be difficult to implement, especially in developing countries. The reported yearly incidence usually ranges from 0.1 to 1 per 100,000 inhabitants in temperate climates and is higher than 10 per 100,000 inhabitants in tropical regions (Haake et al., 2015). The animal reservoir includes mostly rodents, they excrete leptospires in their urine and thus contaminate hydric environment, transmitting the disease to other animals or to humans (Bharti et al., 2003; McBride et al., 2005). Leptospirosis outbreaks emergence are associated with flooding or abundant precipitation (Lacerda et al., 2015; Lopez et al. 2019). Flooding may lead to disruption of health services and damage to households and water and sanitation networks, displacing populations and increasing the risk of exposure to rats and pathogens (Lau et al., 2010). Generally, floods mainly affect the most vulnerable social groups with little knowledge of the risk factors and transmission of the disease (Ricardo et al., 2018).

Northeastern Argentina is severely and recurrently affected by floods, caused by increasingly abundant and intense rains (Lovino et al., 2018a, b). This region accounts for the highest annual number of cases and deaths due to leptospirosis, being a top priority health issue at a regional level (Moral et al. 2014). The National Epidemiological Surveillance System of Argentina (SIVILA) has defined leptospirosis as a notifiable disease (SIVILA 2013). Lopez et al. (2019) analyzed the spatio-temporal variation of leptospirosis for the provinces of Entre Ríos and Santa Fe in northeast Argentina. The incidence of leptospirosis was significantly higher in flooded areas near rivers or in lowlands affected by extreme precipitation. Outbreaks of leptospirosis occurred in months with moderate temperatures (late summer, early autumn) mainly coincident with El Niño event periods, characterized by abundant precipitation and high hydrometric level (close to or above the evacuation level) (López et al., 2016; 2019). The most affected cities were those with the largest population within each province. Although some cities have infrastructure to control floods, leptospirosis outbreaks also appeared during extreme climatic events. A model that facilitates knowing the behavior of the disease considering hydroclimatic variables, it would be a very useful analysis, prediction and prevention tool.

The most classic model is the Susceptible-Infectious-Recovered Epidemiological Model (SIR) that has been used to describe the transmission dynamics of many infectious diseases. The SIR model was initially proposed by Kermack and Mckendrick (1927) and can be modified to adapt it to different infectious diseases for example Dengue (Andraud, et. al, 2012) or the recent COVID-19 (Carcione, et. al, 2020). SIR model divides the human population into three compartments: Susceptible, Infected, and Recovered, then the total human population is defined as the sum of the populations in each compartment. Massad et al. (2011) faced with the evidence of the impact of climate change on the spread of vector-borne infections, highlighted the need to incorporate this component in the epidemiological modeling. In that sense, Triampo et al. (2007) and Pimpunchat et al. (2013) described the transmission dynamics of leptospirosis in Thailand (Asia) through a SIR model where the infection rate varies in relation to precipitation, since in such zone there is a strong rainy season determined by the Monsoon regimes. More recently and for the same region, Chadsuthi et al. (2021) compared 10 different leptospire transmission models, between humans, livestock, and the contaminated environment, also involving flooding and weather conditions. They found that the contact with contaminated environments under flood conditions leads to a higher number of infected individuals.

In the northeast of Argentina, the system is as complex as in Thailand because, besides the precipitation events, there are other hydroclimatic indicators that also would be influencing the rate of transmission disease, as the ONI index and the hydrometric level (Lopez et al., 2019). Thus an epidemiological model such as SIR would be a useful tool to understand the behavior of the disease in a system in which precipitation would not be the only determinant factor for a disease outbreak.

Among the most relevant works about dynamical modelling of leptospirosis it can be mentioned the work of Holt et al. (2006) who propose to model the dynamics of infection with an African rodent (*Mastomys natalensis*) that is thought to be the main source of infection in some regions of Tanzania. The model, representing the climatic conditions in central Tanzania, suggests a strong seasonality in the force of infection on humans with a peak in the abundance of infectious mice between January and April in agricultural environments. The results indicate that removal of animals by trapping rather than reducing the suitability of the environment for rodents will have the greater impact on reducing human cases of leptospirosis. On other hand, Zaman (2010) and Zaman et al. (2012), based on the model of Triampo et al. (2007), presented two-linear models combined of human and vector populations and made a rigorous mathematical analysis of the global stability and optimal control strategies to reduce the proportion of the infected human, in terms of a control variable (antibiotics). In the work published by Sadiq et al. (2014) the authors consider a leptospirosis epidemic model with nonlinear saturated incidence by applying the optimal control techniques to eradicate the infection in the human population. In order to find such optimal control techniques for the eradication of leptospira in the host population, they define three control variables, one for humans and the second and third one for the vector population. Except for the works of Triampo et al. (2007), Zaman et al. (2010 and 2012) and the recent one of Chadsuthi et al. (2021), the others did not calibrate their models against observed or registered data, they only did practical mathematical exercises based on physical-biological hypotheses of the interaction between the vector and the susceptible population.

According to our background review, in South America, and specifically in the study region, there is no scientific literature on the implementation of this type of models combined with hydroclimatic variables (López et al., 2018). However, it could be mentioned the recent work of Gualtieri and Hecht (2019) who designed a simple deterministic model based on differential equations with promising results, but only explored the model dynamics by computational simulations without testing it with registered data.

In this paper, we proposed a model for leptospirosis outbreaks using the classical SIR Epidemiological Model but incorporating hydroclimatic variables that influence the disease transmission in the northeast of Argentina, fitting the model to the actual data reported in outbreak events. The incorporation of these variables into a SIR model will enable us to better understand the behavior of the disease in outbreak events by making a contribution that may improve public health policies in the region. The model is implemented in the three most populated cities in the area, Santa Fe, Rosario and Paraná, which have latitudinal and topographic differences. Each of the cities has experienced at least one outbreak in a decade of reported cases (2009-2018).

The paper is structured, first with the introduction to the problem, then section 2 presents the datasets and the methodology used: the preparation of two functions that incorporate hydroclimatic variables into the SIR model, the structure of the model and a sensitivity analysis. Section 3 presents the results and discussion of the previous analysis, the model implementation and the sensitivity analysis. Finally, section 4 concludes about the numerical results of the model and its potential as a prediction tool.

## 2. Materials and methods

### Study area

The study area is shown in Figure 1. It spans over the Santa Fe and Entre Ríos provinces located in the northeast of Argentina, southeastern South America (Figure 1a). The region is an extended plain crossed by numerous watercourses (Figure 1b). The predominant climate is temperate with hot summers and no dry season, according to Köppen-Geiger’s climate classification (Peel et al., 2007).

**Figure 1:**
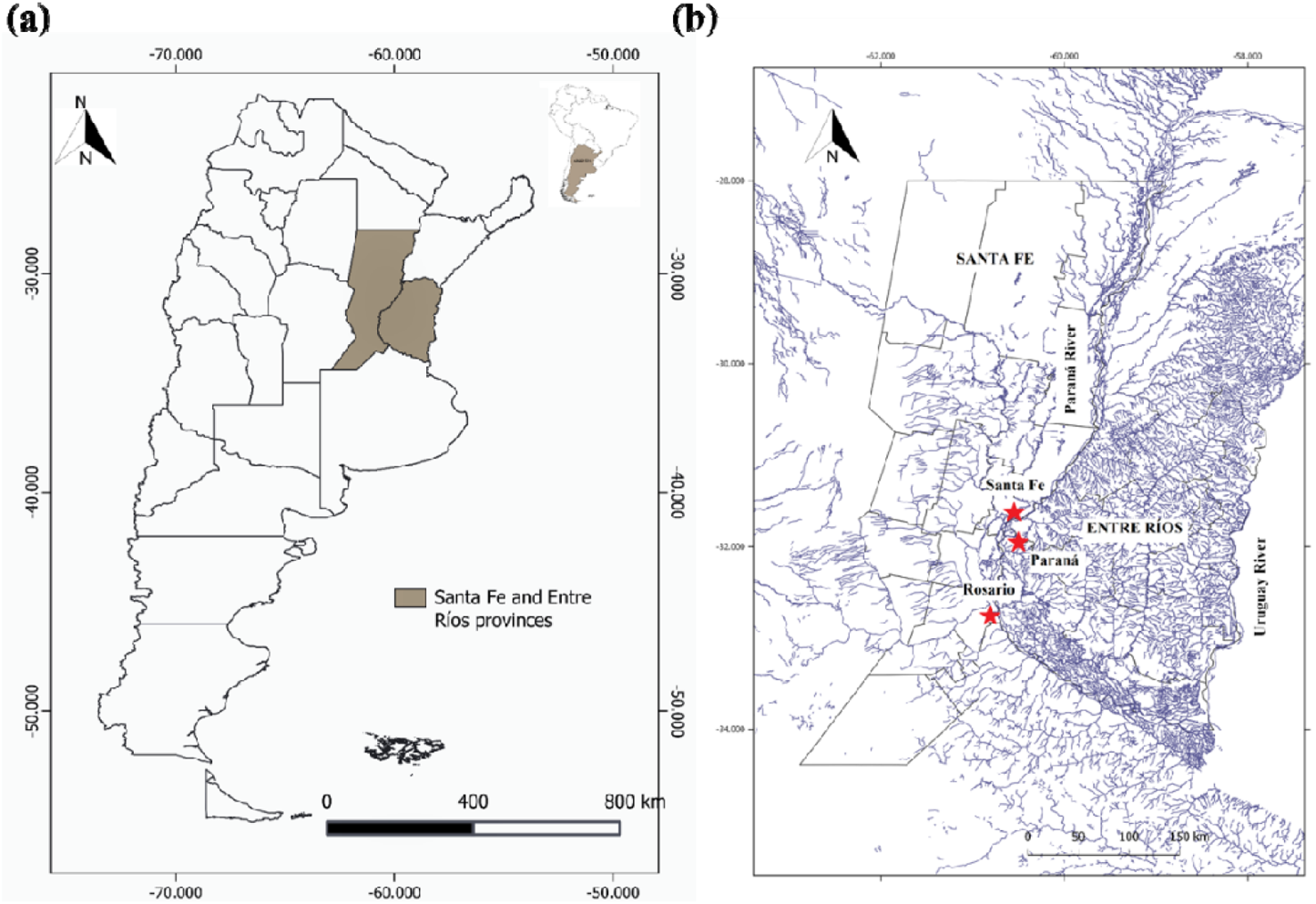
Study area in Argentina. (a) Santa Fe and Entre Ríos provinces. (b) Hydrography of the area and location of the studied cities in red.

The Paraná River is one of the most important rivers of South America and the main one in northeastern Argentina (Fig. 1b). The upper and middle portions of the Paraná River have a maximum river level in late austral summer. The upper Paraná basin (south of Brazil) provides the largest amount of water to the Paraná River flow. The dynamic of the Middle Paraná River floodplain is strongly shaped by cycles of water level rises and falls, and due to its flat landscape, it is highly susceptible to flooding due to river overflows following heavy precipitation (Drago, 2007). Thus, the Paraná floods in northeastern Argentina are a direct consequence of excess precipitation in the upper Paraná Basin that has a close link with El Niño events (Berri et al., 2002a). The Salado river is also an important one, particularly in the study area, that crosses the north of Argentina from northwest to southeast (Fig. 1b). This river flows into a secondary channel of the Paraná River in the city of Santa Fe (capital of the Santa Fe province).

The three most important cities of the two provinces are located on the river banks: Santa Fe and Rosario (Santa Fe province) and Paraná (Entre Rios province), indicated in Figure 1b. The city of Santa Fe has a flat riverine topography and the Paraná and Salado rivers influence it. This city is protected in its limits by defenses that avoid flooding by the surrounding rivers. It also has electric pumps that evacuate excess water from fluvial or rainy floods. Rosario city has a flat topography but is located higher above sea level than Santa Fe city and over the coast of the Paraná River presents a ravine geomorphology. Paraná city presents hills that condition the water runoff and, like Rosario, has ravine topography on the Paraná River coast. Santa Fe and Paraná are only separated by the Paraná River, at the same latitude, while the city of Rosario is located about 180 km south from the other two cities (Fig. 1b).

The location of these cities makes them highly vulnerable to flood episodes or extreme precipitation events (Lovino et al. 2018c). Additionally, the migration of people from smaller cities or the countryside to larger cities in search of better working conditions also increases the vulnerability. This migration of people causes the occupation of non-habitable lands, near rivers or in low-lying areas, increasing poverty and vulnerability (Alberto, 2012). A study carried out in the city of Santa Fe reveals that 22% of the population presented high vulnerability to floods and a 27.88% very high socio-environmental vulnerability (Gómez, 2007). This is mainly due to the occupation of non-urban land in the flood valley of the Salado river and the lack of urban services (Cardoso, 2019). In the case of Rosario city, the areas of high vulnerability are located towards the periphery of the city, with some exceptions in the interior of the city that are generally a consequence of irregular population settlements. The potential for disasters increases due to the combination of such environmental conditions with the large population with high vulnerability (Ruiz et al., 2019).

### Hidroclimatic Datasets

Hydroclimatic datasets include in-situ monthly total precipitation, monthly mean temperature, maximum monthly hydrometric level and the Oceanic Niño Index (ONI, NOAA/NWS/CPC). Precipitation and temperature data were provided by the National Meteorological Service (SMN) of Argentina. The meteorological stations include Sauce Viejo Aero (near the city of Santa Fe), Rosario Aero and Paraná Aero. The National Water Institute (INA) of Argentina provided hydrometric data, while the Argentine Naval Prefecture (PNA) provided river evacuation level data. The ONI Index is used to determine the years and months under El Niño, La Niña or Neutral conditions, and is defined as the 3-month running mean SST anomaly for the Niño 3.4 region (5°N-5°S, 120°-170°W, https://ggweather.com/enso/oni.htm).

### Epidemics Datasets

This study is carried out for the cities of Santa Fe, Paraná and Rosario that reported leptospirosis outbreaks in recent years. The leptospirosis epidemics are documented between 2009 and 2018 (no data yet available for 2019 and 2020). National System of Epidemiological Surveillance by Laboratories of Argentina (SIVILA) was implemented in 2009, and since then the notification of leptospirosis is mandatory in Argentina. Moral et al. (2014) provide guidelines for the confirmation of leptospirosis cases in Argentina which is carried out by: (a) a positive MAT (microscopic agglutination test), bacterial isolation and detection of bacterial genome by PCR1 (Plant Cadmium Resistance protein), (b) MAT seroconversion in two or more samples, preferably with an evolution of more than 10 days, (c) verification of exposure to the same source and at the same time as a confirmed case (a) or (b). The Ministry of Health of each province analyzes the laboratory, clinical and epidemiology information and makes the classification according to the confirmation criteria. We assess only the confirmed cases according to the information provided by the Directorate for Health Promotion and Prevention, Ministry of Health of the Santa Fe province and the Epidemiology Division of the Entre Ríos province. The total number of confirmed leptospirosis cases between 2009 and 2018 was 263; 94 for Santa Fe, 97 for Rosario and 72 for Paraná. This study does not analyze probable cases neither nor suspected or unconfirmed cases.

Data about population indicators were retrieved from the 2010 National Population, Household and Housing Census, National Statistics and Census Institute (INDEC) of Argentina.

### Methodology approach

A series of previous analyses was performed to define the parameters that would be incorporated into the epidemiological model.

Following the methodology used in Lopez et al. (2019), a Principal Component Analysis (PCA) was carried out to determine the hydroclimatic indicators that influence the presence of outbreaks in the three cities and that would finally be explicitly incorporated into the model. The hydroclimatic indicators considered in PCA were monthly total precipitation, maximum monthly hydrometric river level, the ONI; and the number of cases is the variable used to quantify the magnitude of the outbreak in the 2009-2018 period.

In order to determine the function between the hydrometric level and the flooded area in each city, Landsat images TM5 and 8/OLI (approximate error of 30 m, one pixel) were analyzed using the Quantum Gis software. A buffer of 6 km^2^ was defined around the hydrometer of each city to calculate the flooded area in three situations: minimum hydrometric level (year 2009), average hydrometric level (year 2015) and maximum hydrometric level (year 2016). Therefore, three Landsat images per city were analyzed whose scenes are 227-082 path-row for Santa Fe and Paraná cities and 228-083 path-row for Rosario city. Figure 2 presents the satellite products used in the analysis. Figures 2a, 2b, 2c show the satellite images of the cities of Santa Fe and Paraná, and figures 2d, 2e, 2f, the images for Rosario city, that represent, from left to right, the low, medium and high hydrometric level, respectively. The images were processed in the reflectivity values and the applied atmospheric correction was Dark Object Subtraction (DOS) method (Chavez, 1988). The modified normalized difference water index (MNDWI) has been successfully used to delineate surface water features (Xu, 2006). From the Landsat imagery, a binary water mask was created using the MNDWI in which any negative value was classified as water (Xu, 2006). The MNDWI of Landsat data is defined by:

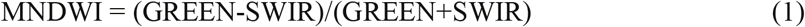

where GREEN is the radiance at green wavelengths (0.52–0.60 μm) and SWIR is the radiance at the Short-wavelength infrared (1.55–1.75 μm). Once the flooded areas were obtained according to each hydrometric level (*h*), the mathematical function of the variation of the flooded area (**Δ**FA) was determined for each city (**Δ**FA = *f*(*h,t*)). At difference of Chadsuthi et al. (2021) who counted the number of flooded pixels to calculate an index of land flooding, here a function that relates hydrometric level and flooded area is obtained and it is directly incorporated to the model.

**Figure 2:**
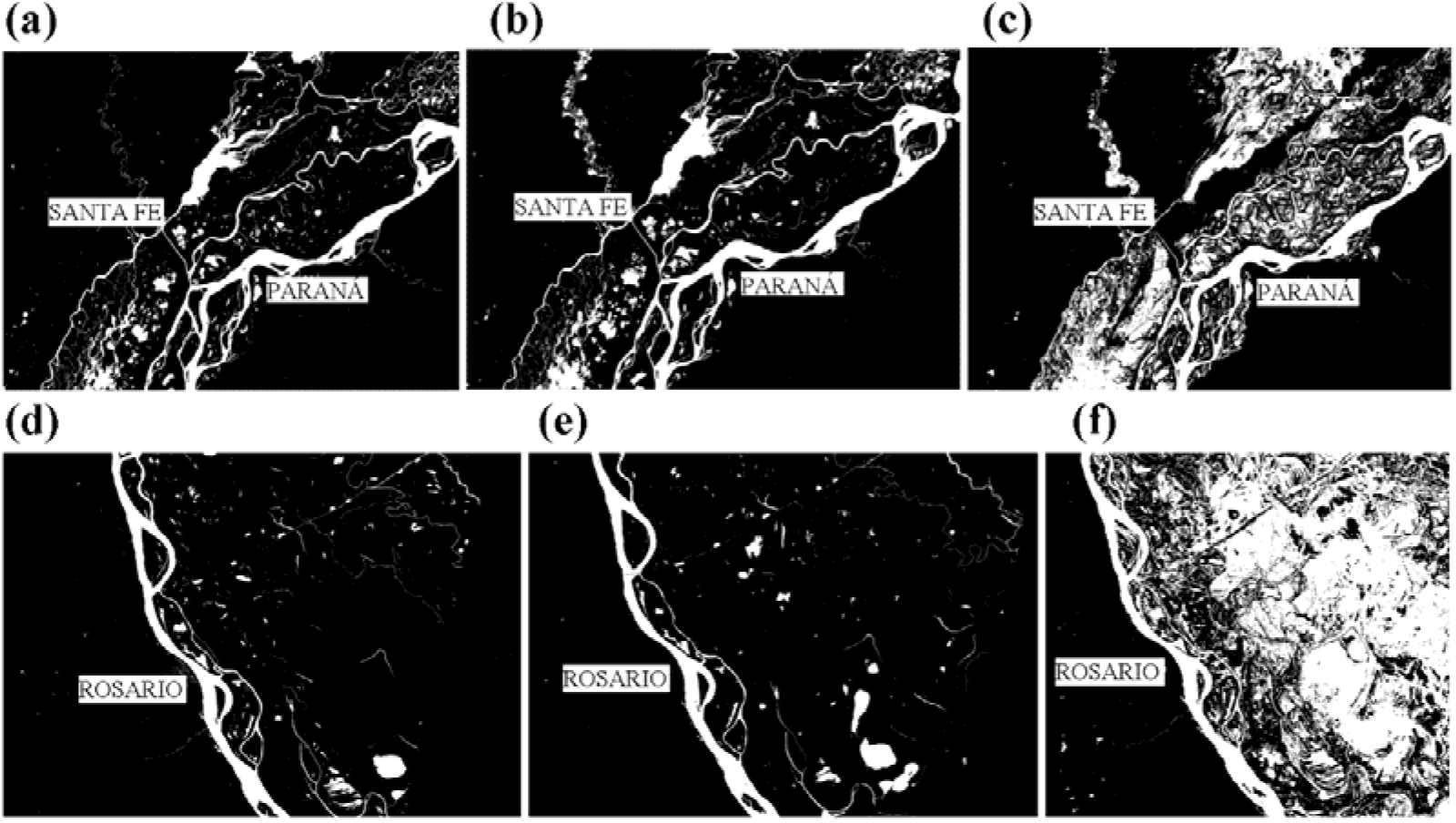
Landsat images TM5 and 8OLI of the study area used to build the FA function. The areas occupied by water are white; areas of dry land are black. a) and d) year 2009, minimum hydrometric level; b) and e) year 2015, average hydrometric level; c) and f) year 2016, maximum hydrometric level. Upper row (a, b and c) around the cities of Santa Fe and Paraná. Lower row (d, e and f) around Rosario city.

Finally, based on the methodology of Triampo et al. (2007), the monthly distribution of precipitation of each year is modelled with a Gamma probability distribution function and incorporated into the SIR model. The distribution is defined using non-linear least squares to fit the function to data of precipitation, where t is the time, is the shape parameter, and *loc* is the location parameter.

### Epidemiological Model

The adopted model is based on the work of Triampo et al. (2007), which considers a deterministic model for the transmission of leptospirosis in the Thai population. As many classical SIR models, it takes into account the following hypotheses: *(a)* the total size number of human population is constant; *(b)* the natural death constant rate λ_H_ is taken to be the same for all population subgroups; *(c)* the individuals are unaffected by age or disease status so that the vital statistics of all individuals are the same, the life expectancy is the same for everyone and is 1/ λ_H_ ; (d) deaths are balanced by births (birth rate being μ_H_) and *(e)* all newborn are considered not to be immunized and so become vulnerable instantly. In addition, there are a few assumptions in particular to the spread of leptospirosis such as: *(a)* Only infected vectors can be infected human, this means that an infected human cannot infected another human; *(b)* infected humans cannot infect the susceptible vectors: *(c)* once infected, a susceptible vector becomes instantly infectious with no incubation time needed for the infectious agents to develop; *(d)* the infected human can be cured by the antibiotic medicines and they become immune at a rate r_1_ and immune individuals (humans) become susceptible again at a constant rate r_2_; *(e)* As a difference of Triampo’s model, this considers the rate of transmission of leptospirosis from an infected vector to a susceptible human varies with the amount of rainfall that follows a gamma distribution (*Γ*) and the function of variation of the flooded area (**Δ**FA); (f) *Γ* and **Δ**FA modulate the strength of infection assuming that they increase or decrease the probability of an encounter between a vector and a human host. Figure 3 shows the diagram of the proposed model.

**Figure 3:**
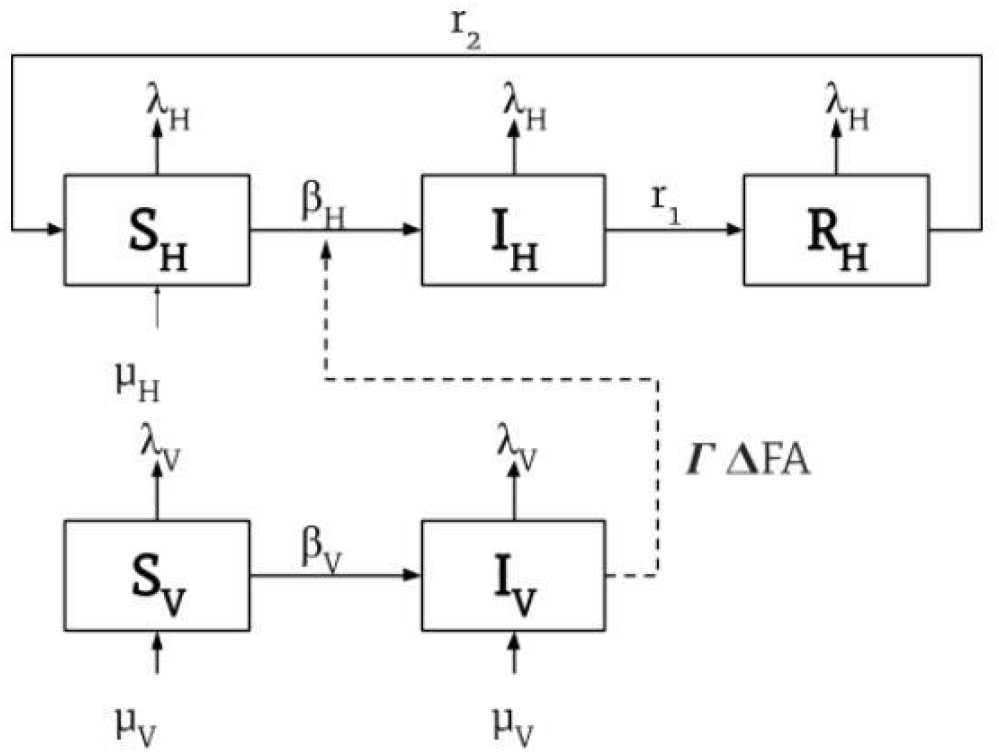
Model description. *S*_*H*_ (Susceptible Human), *I*_*H*_ (Infected Human), *R*_*H*_ (Recovered Human); *S*_*V*_ (Susceptible vector), *I*_*V*_ (Infected Vector). Adapted from Triampo et al. (2007).

The Ordinary Differential Equations (ODE) system that describes the dynamic SIR model is as follows:

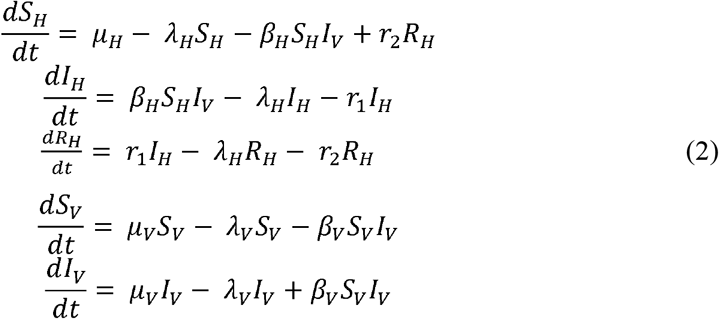

The model parameters are *μ*_*H*_ human birth rate; *λ*_*H*_ human death rate; *β*_*H*_ infection rate, *r*_1_human antibiotics recovery rate; *r*_2_ human immunity loss rate; *μ*_*V*_ vector birth rate; *λ*_*V*_ vector death rate and *β*_*V*_ vector infection rate. Infection rate *β*_*H*_ is non constant in time and is defined as a function of ***Γ*** and **Δ**FA as:

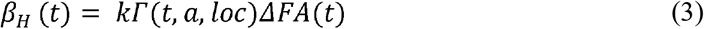

where *k* is a scale parameter.

The ODE system (2) is implemented on a python code, solved with the *my_ls_func* function from the scipy library, and it is fitted using a nonlinear dog-box method with the least squares function (SciPy Reference Guide, Release 1.4.1).

Normalizing each run of the model, the initial conditions of the ODE system were set up as S_H_(0)=1, I_H_(0)=0; R_H_(0)=0; S_V_(0)=1-1×10^−2^ and I_V_=1×10^−2^.

The model parameters were set up considering the model hypothesis mentioned in the methods section. The life expectancy of a human (1/*λ*_*H*_) is about 75 years old and so the mortality rate of the human (*λ*_*H*_) is 1/(365 x 75) per day. This value is slightly higher than reported from Triampo et. al (2007) since it was updated and adapted to the population of the study region. The life span under natural conditions of the vectors (rats) is 1.5 years. Therefore, the death rate of the vectors (*λ*_*V*_) is 1/(1.5 x 365) per day. The *β*_*V*_ vector infection rate, recovery rate of an infectious human, or immunity (*r*_1_) and the rate of loss of immunity (*r*_2_) are parameters that were adjusted by calibrating the model to the actual data in each city. Infection rate *β*_*H*_ was calculated as previously explained, incorporating hydroclimatic variables, by means of ***Γ*** and **Δ**FA functions.

The Root Mean Square Error is used to compare observed and modeled data, according to equation (4), where *x* is the number of leptospirosis cases at time *i*, and *n* is the total number of time steps at which the model is evaluated.

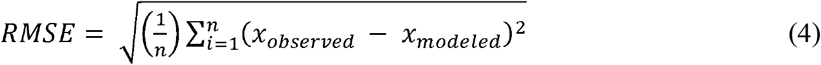

The RMSE is always positive and has the same units as the variable, so is a good measure of the goodness of fit to real values, with the smaller RMSE the better fit of the model.

### Sensitivity Analysis

In relatively complex models such as the one presented here, sensitivity analysis provides confidence about its general behavior when the input parameters are modified one at a time. The objectives of the sensitivity analysis can be several, for example define model adjustment sequence, focusing on those parameters that cause more sensitivity in the model results, or to provide a range of uncertainty both for the actual values and physical meaning of the parameters involved as well as in the results obtained, among others (Pianosi et al., 2016).

A sensitivity analysis of the model results in a change of the most relevant parameters was performed using the data of Santa Fe city for year 2010 as a benchmark case. The corresponding effect on modeled leptospirosis cases was analyzed varying the infection rate between humans and vectors and the proportion of infected vectors population, which are the two relevant parameters that control the outbreak evolution.

## 3. Results and discussion

Prior to advance in the discussion of the model results, Figure 4 shows the time series of registered leptospirosis cases for the 2009-2018 period in the three selected cities. Four marked outbreaks events appeared in 2010, 2013, 2014 and 2015 in the three cities, being the 2010 the largest one. In the three cities, the data during outbreak years show a seasonality that occurs in late summer and early fall (between January and March), with the main peaks in cases in February, as already analyzed by López et. al (2019). It should be noted that the city of Santa Fe, despite being 2.5 times smaller than the city of Rosario, during outbreaks presented total values of leptospirosis cases similar to the latter.

**Figure 4:**
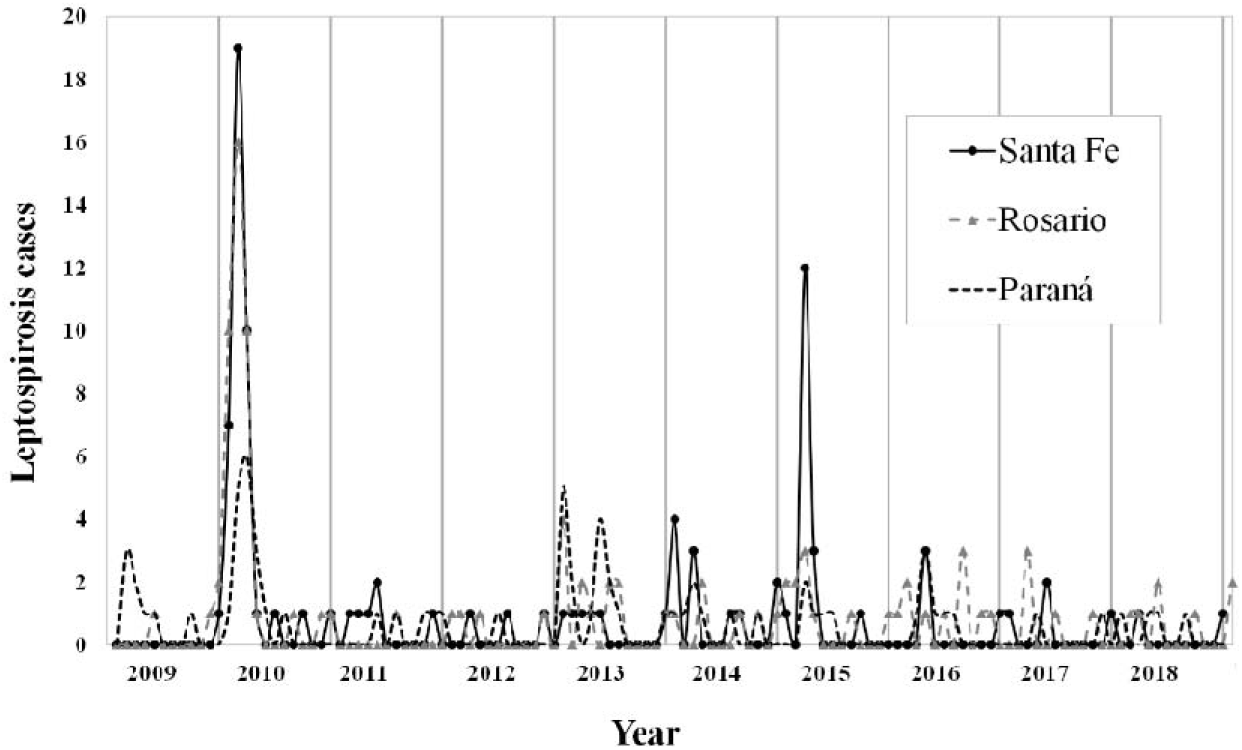
Leptospirosis cases for the period 2009-2018 in Santa Fe, Rosario and Paraná cities (northeast of Argentina).

### Pre-model analysis

To determine if the selected hydroclimatic indicators are correlated with the presence of an outbreak, a PCA analysis was performed. The results are shown in Figure 5 and the data in Table 1. In the figure it can be observed how the three cities with outbreaks are grouped to the right of the biplot and correlate with the analyzed indicators. i.e., monthly total precipitation, maximum monthly hydrometric level of the river and ONI. Therefore, all the hydroclimatic indicators evaluated are highly correlated with the leptospirosis outbreaks in the studied cities, with a coefficient of correlation of 0.998. In this sense, an integrating analysis of the hydrometric indicators is necessary given the complexity of the system in the northeast of Argentina.

**Figure 5:**
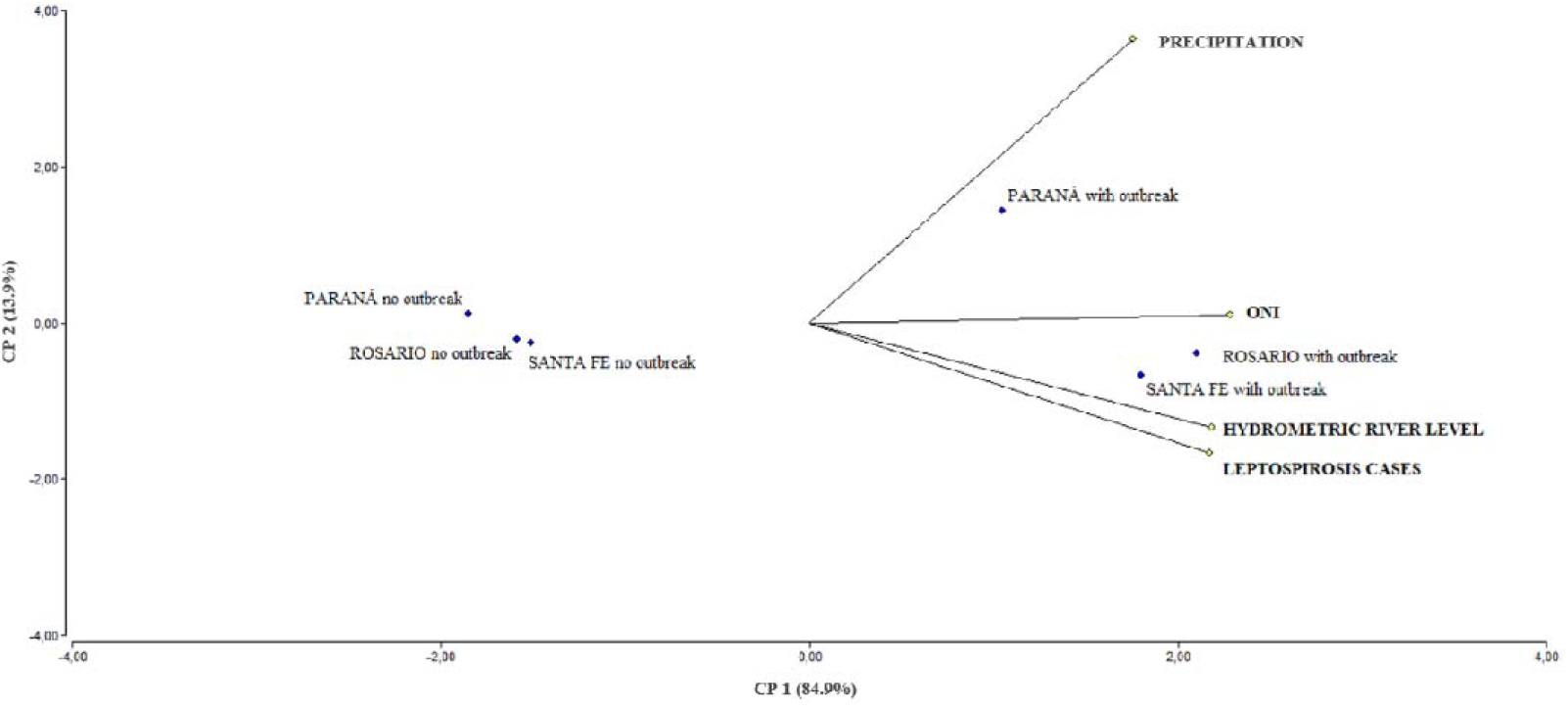
Principal Components Analysis biplot that correlates the outbreaks of leptospirosis in the cities of Santa Fe, Rosario and Paraná with the hydroclimatic variables including monthly mean precipitation, maximum monthly hydrometric level and ONI index.

**Table 1:** Correlation with the original variables

| Variable | CP 1 | CP 2 |
| --- | --- | --- |
| Precipitation | 0.76 | 0.63 |
| Leptospirosis cases | 0.90 | -0.41 |
| Hydrometric river level | 0.90 | -0.39 |
| ONI | 0.95 | 0.25 |

It must be mentioned that ONI is not completely independent of the rest of the analyzed variables since in this region during El Niño years, precipitation is higher than normal, as well as the hydrometric level (Berri et al., 2002b). Therefore despite the index correlates with the leptospirosis outbreaks is not considered at this stage of the modeling in order to obtain a model with few variables, which simplifies the results interpretation.

Additionally, the variation of the flooded area was analyzed according to the hydrometric level and, as expected, it increases with increasing hydrometric levels, but differently for each of the cities. Table 2 shows the functions and their corresponding parameters for each of the cities. In the case of Santa Fe city, a linear function was obtained, while for Paraná and Rosario cities, a quadratic relationship between hydrometric river level and flooded area was obtained. It is likely that the topography of the cities is determining these functions. Paraná and Rosario have ravines on their coasts of the Paraná River, while Santa Fe presents a flat topography with a smooth topographic gradient towards the alluvial plain of the Paraná River, which could be the cause of the different relationship structure between the hydrometric level and the flooded area in each city.

**Table 2:** **Δ**FA functions for the three selected cities.

| City | $\Delta FA$ function | Parameters |
| --- | --- | --- |
| Santa Fe | $\Delta FA = c_1 h + c_2$ | $c_1 = 85.828; c_2 = 1531.5$ |
| Paraná | $\Delta FA = c_1 h^2 + c_2 h + c_3$ | $c_1 = 218.71; c_2 = -1345.4; c_3 = 4095.1$ |
| Rosario | $\Delta FA = c_1 h^2 + c_2 h + c_3$ | $c_1 = 679.16; c_2 = -3320.1; c_3 = 7851.4$ |

As hydrometric river level varies with time, *h(t)*, **Δ**FA is **Δ**FA*(t)* as well, and each function is calculated and incorporated in the model in each time step, once the hydrometric level is known. As an example, Figure 6 shows the **Δ**FA functions throughout a particular year, 2010, for each of the three cities, calculated for the time steps of the model and using the hydrometric Paraná River level data. The year 2010 is selected because it is one of the years when disease outbreaks occur in the three cities, as it was mentioned in the previous section. The figure shows a linear behavior of the function for Santa Fe city, and a quadratic one for the other two cities.

**Figure 6:**
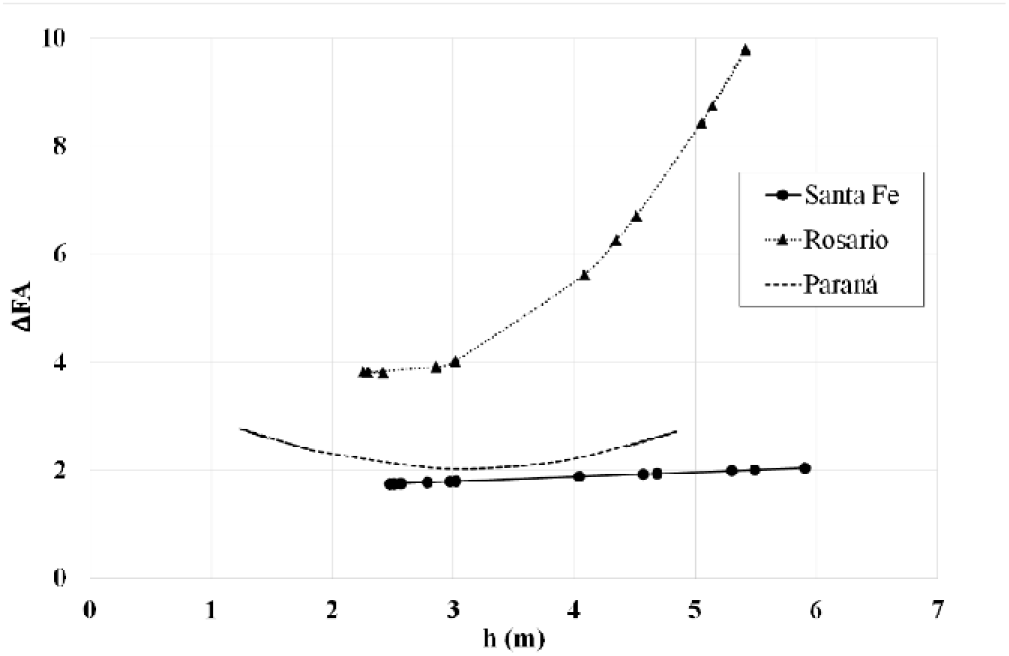
FA functions for each city according to hydrometric Paraná River levels registered for 2010.

In summary, the flooded area function considers the orographic component since it relates hydrometric river levels with water occupied areas and behaves linearly over smooth coasts and quadratically in ravine environments.

In the case of the distribution the procedure is similar. First, it is adjusted to the precipitation data in order to obtain the parameters, and then the function is incorporated into the SIR model. Figure 7 shows the normalized precipitation data of Santa Fe city in 2010 and its Gaussian adjustment.

**Figure 7:**
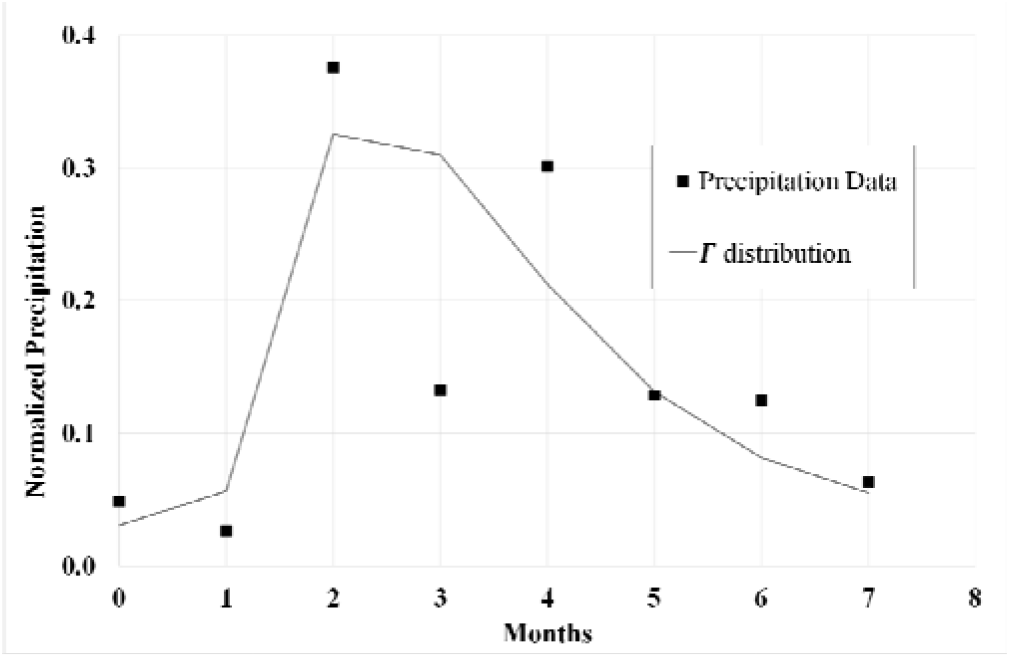
Normalized precipitation for Santa Fe city for 2010, distribution adjusted. Parameters: =2.47 and *loc*= 0.9

### Model results

As it was mentioned before, the time series of leptospirosis cases shows an important outbreak in 2010 in the three cities (López et. al, 2019), so that the model is evaluated in that particular year. Figure 8 shows registered cases of leptospirosis in the three cities, as well as the SIR model results for 2010. The Santa Fe city outbreak lasted only three months, from January to March 2010, with a maximum peak of 19 cases in February and 40 confirmed cases in total. In the same year, Rosario city presented 40 confirmed cases, also concentrated between January and March with its peak of 16 cases in February, and Paraná city had 17 cases in total with an outbreak peak of 6 cases in March. In the three cities it can be observed some isolated cases during the winter months which may be associated with the specific activities of affected people (rural environment or in contact with contaminated water). The number of cases of leptospirosis infections obtained in the three cities by solving the proposed SIR model for 2010 are in good agreement with the actual data. The model is able to reproduce the peak behavior in Santa Fe, Rosario and Paraná, as can be observed in Figure 8a, 8b and 8c respectively.

**Figure 8:**
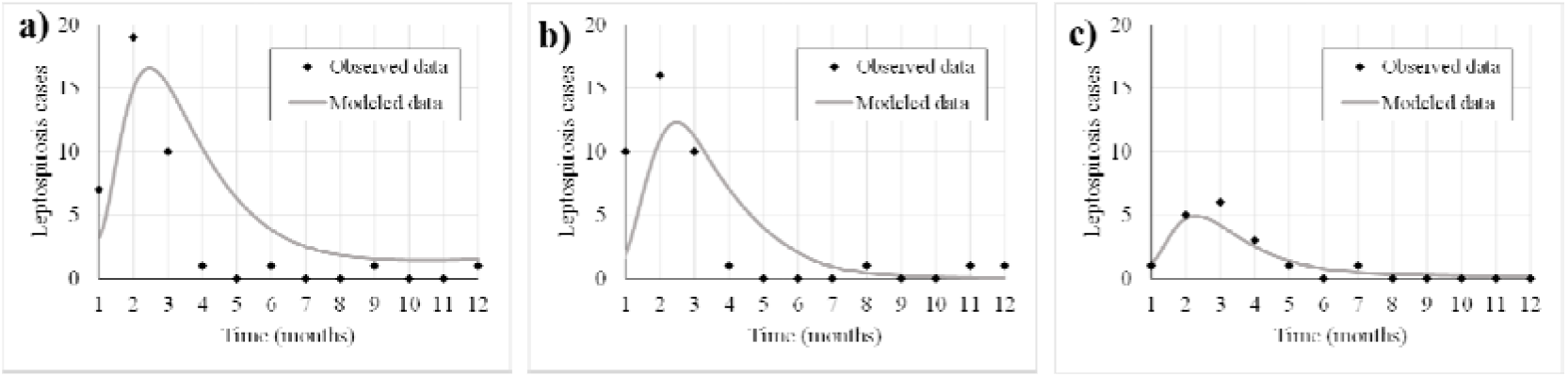
SIR model results and registered cases for the leptospirosis outbreaks in 2010 in a) Santa Fe, b) Rosario and c) Paraná.

Calculated RMSE are 4.1; 3.5 and 0.6 cases, for Santa Fe, Rosario and Paraná cities, respectively. The higher values in the first two cases may be due to the fact that the number of cases at the peak of the outbreak is relatively greater and the model needs a longer tail to decrease that peak, so that it overestimates the number of cases during the subsequent months.

The calibrated value of the parameters cannot be directly compared with literature since the structure of models of other authors has been developed for different purposes so that it is slightly different from the one presented here. However, works of Zaman (2010; 2012); Minter et. al (2018) or Gualtieri and Hetch (2019) have obtained similar results under equivalent interpretations of the physical meaning of the parameters.

The last aspect to point out is that, in years when a disease outbreak is not evident, the model does not perform very well. To exemplify this, Figure 9 shows the results for Santa Fe city in every year of the study period. In the outbreaks of 2010, 2014 and 2015, the model reproduces the observed data with RMSE of 4.1, 1.5 and 2.6, respectively. Instead, in the rest of the years when there are only isolated cases that do not show an outbreak wave, the model reaches the amount of cases in the first months of the year forced by the function of rainfall and flooded areas, but it does not capture the isolated cases of the following months. The low RMSE values in those years is due to the low number of cases that occur, unlike the years mentioned with outbreaks (2010, 2014 and 2015). This aspect can be a consequence of the model structure that assumes a higher probability of an outbreak when it detects an increase in precipitation and flooded areas. However, when these variables are not increased, the model cannot detect the increase in the number of cases.

**Figure 9:**
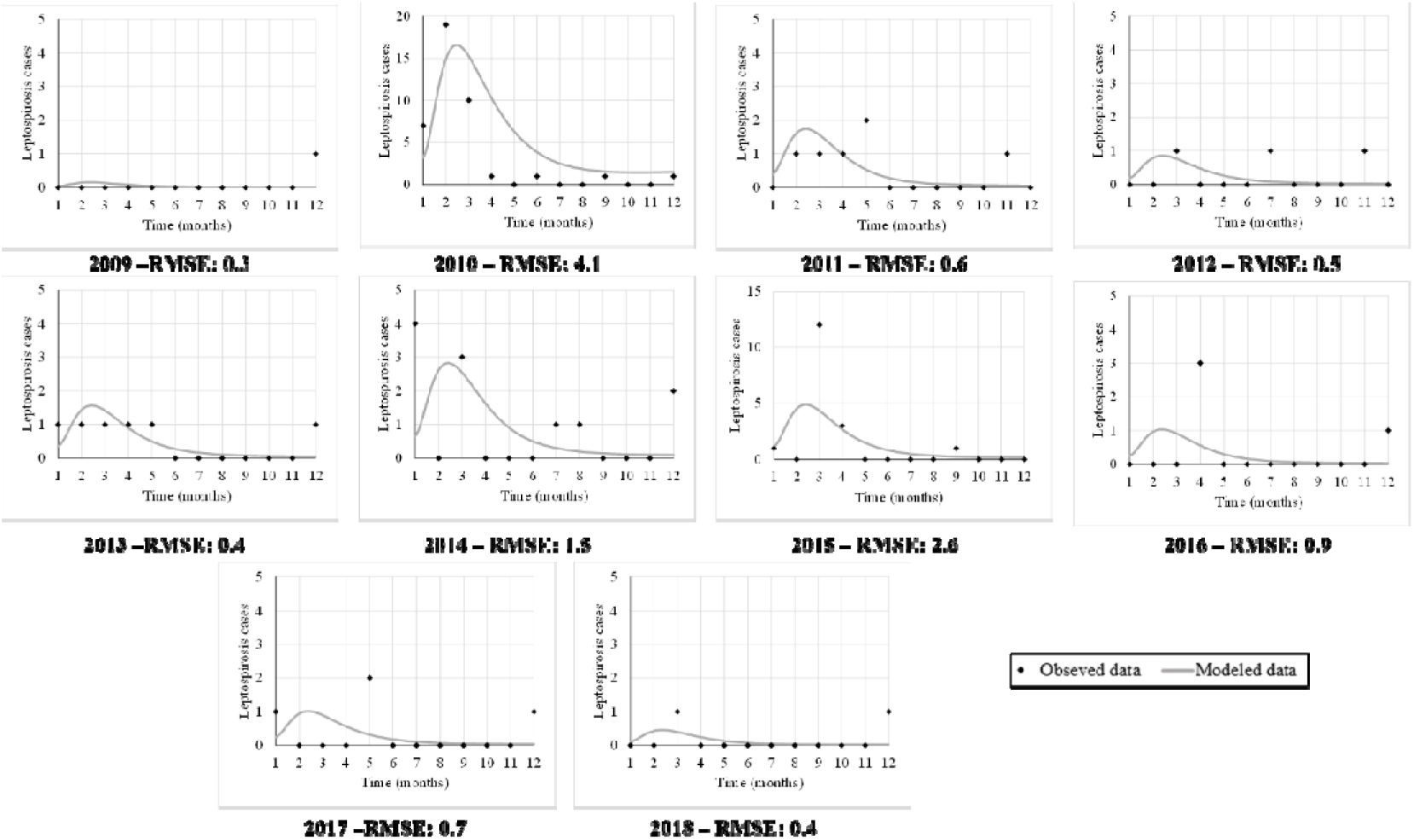
Model results of leptospirosis cases for Santa Fe city per year of the study period vs. observed data. Note: 2010 and 2015 vertical axis show a different scale due to the increase in cases during these outbreaks.

These results are in concordance with Chadsuthi et al. (2021) who pointed out that when the numbers of cases is low or medium the analysis cannot provide a significant result with determinist models like the one used in this work. Nevertheless, it remains valid in years with obvious outbreaks which constitutes an advance in the modeling of this infectious disease in the region.

The non-appearance of an outbreak in the most of years (except from 2010, 2014 and 2015) could be due to environmental conditions were not the precursors to outbreaks or because of social factors such as prevention or prophylaxis measures that are not explicitly considered in the model, but are issues to be incorporated into future work. Following the findings of Chadsuthi et al. (2021) another hydroclimatic variable to explore in next work is temperature. They concluded that including the effect of temperature improved their transmission model by only a modest amount, but in a climate change context like the present one, when the temperatures are steadily increasing, its possible effect on the incidence of leptospirosis should not be underestimated and a modeling application can be very useful to evaluate it.

In summary, as a next step it would be important to evaluate two additional variables: one that would be reducing them in years that outbreaks would be expected (prophylaxis) and another that would apparently influence the increase in cases (temperature).

### Sensitivity Analysis

The infection rate *β*_*H*_ (eq. 3) indicates the probability of transmission when a susceptible human is exposed to the infectious agent through contaminated water, by either precipitations or floods, and arises as a combination of both ***Γ*** and **Δ**FA functions. Those functions are obtained independently from the model, as explained before, and cannot be modified. However, it is possible to test the influence of the scaling parameter *k* in the transmission rate, by increasing and decreasing it in 10 and 25 % from its calibrated value. The variation of modeled data results for Santa Fe city due to changes in parameter *k* is shown in Figure 10a and reveal a proportional behavior, as expected. When *k* increases, the transmission rate increases proportionally, while a drop in *k* causes a decrease in the peak of the outbreak.

**Figure 10:**
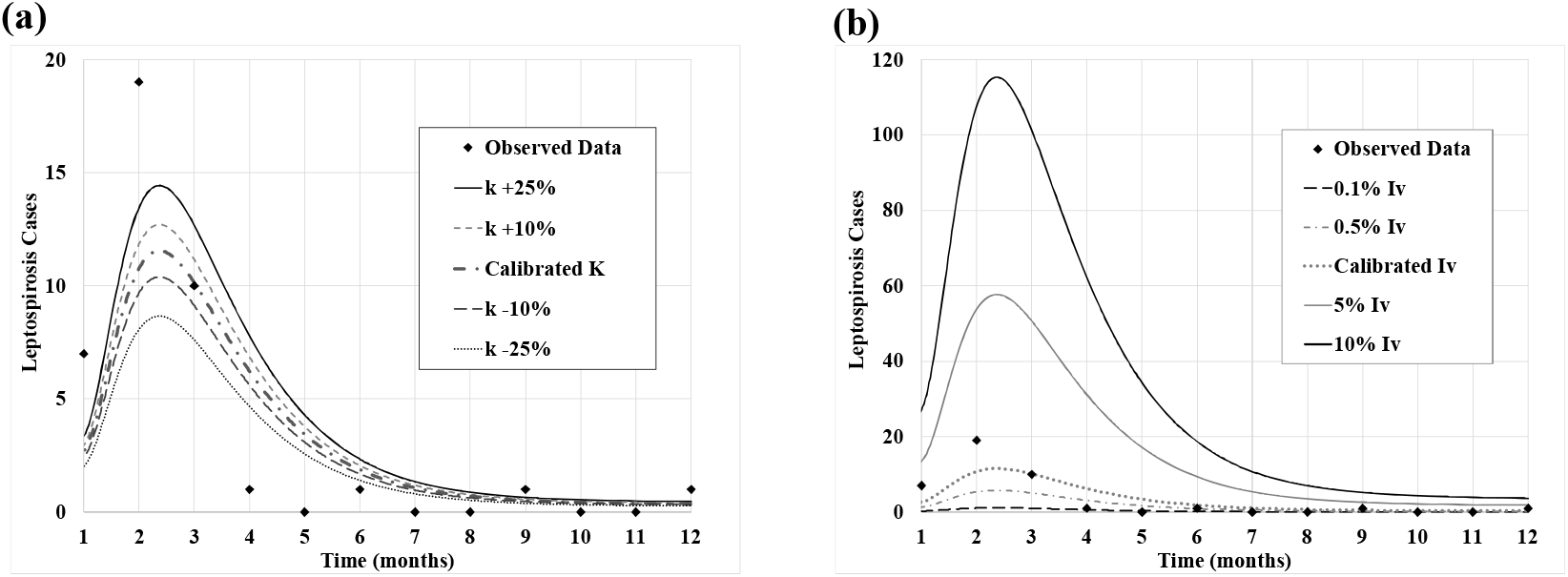
Variation of modeled data results in Santa Fe city, year 2010, when is modified: (a) the scale parameter k (b) the proportion of infected vectors population, *Iv*.

Although apparently the curve that increases the value of *k* by 25% fits better the peak of the outbreak, the RMSE for all the simulated time steps is 2.8, in the same order of magnitude of the calibrated dataset. This is because in the months of the year when there are no cases, the model with *k* increased by 25% overestimates the number of cases.

The k parameter evaluated is equivalent to the *a* transmission parameter mentioned in Gualtieri and Hecht (2019) and both present similar behavior. Similarly, the sensitivity analysis performed by Chadsuthi et al. (2021) also shows that the most relevant parameter is the transmission rate of leptospires to humans, even though they use different model structures.

Parameter *k*, and globally *β*_*H*_both control the disease evolution, so in terms of public health policies, control measures that would decrease these factors, could help in preventing the outbreaks occurrence. Such measures will be mainly diminishing the contact of susceptible populations with contaminated water.

The other parameter analyzed is the proportion of infected vectors population, *Iv*, which acts as an initial condition in the simulations. The tested values were 0.1%, 0.5%. 5% and 10% of the vector population that are infected with leptospirosis at time = 0. Figure 10b shows the variation of modeled data results when the percentage of infected vector population is modified, comparing them with the calibrated value of 1% of *Iv* It can be observed that the model is very sensitive to this parameter, but values of *Iv* greater than 1% yield unrealistic values of leptospirosis cases. In this case, the sensitivity analysis was useful to establish an upper limit of the parameter.

Regarding the actual value to be used in the model, a rodent population study could be performed to quantify what percentage is actually infected, as the one carried out by Ricardo et al. (2020). On the other hand, controlling such population and minimizing the contact with the human population would be effective measures to reduce the impact of a leptospirosis outbreak, just to mention an example.

## 4. Conclusions

The development of a model like the proposed in this paper could be a public health management tool to improve the preparation for eventual leptospirosis outbreaks.

The influence of climatic variables, such as precipitation and the extent of flooded areas are important variables that directly affect the infection rate of the susceptible human population. The extent of flooded areas, caused by the increase of hydrometric levels, obtained from remote sensing, constitutes a very useful tool to analyze its impact at regional scales.

The dynamic modeling of infectious diseases considering hydroclimatic variables constitutes a climatic service for the public health system, not yet available in Argentina.

Since climatic conditions vary in each region, its relevance on the transmission of leptospirosis should be evaluated for each one and this work aims to contribute in that direction.

The number of cases of leptospirosis infections in the three selected cities of the northeast of Argentina, Santa Fe, Rosario and Paraná, obtained by solving the proposed SIR model for the 2010 outbreaks, are in good agreement with the actual data. The SIR model captures the dynamics of the leptospirosis outbreak wave.

The flooded area function considers the orographic component since it relates hydrometric levels with water occupied areas and behaves linearly when there are smooth coasts and quadratically in ravines environments. Knowing this relationship and the future predictions of precipitation, the transmission rate *β*_*H*_ could be estimated and incorporated in the model to predict the probable number of infected humans in advance.

Although the modeling has yielded good results for the 2010 outbreak in the three cities, it is important to consider the underestimation of the disease incidence and the unreliability of the registered data because leptospirosis presents a symptomatology similar to other diseases.

Despite the ability of the model to reproduce the 2010 outbreak, it does not perform very well when isolated cases appear outside the outbreak periods, probably due to factors not explicitly considered in the model. In forthcoming work we plan to generalize the model making it more realistic by including, for example, the time delay between the appearance of symptoms and the confirmation of an infected case. Prophylaxis related variables could also be added in order to explore epidemic control scenarios, like pharmaceutical or non-pharmaceutical implementations.

Other improvements to the model could include gradually changing the structure of the ODE system in terms of the precipitation or hydrometric level functions by testing other formulations, or explicitly including the ONI variable and temperature. Through optimization and control techniques, different disease outbreak control strategies can be tested numerically, which could be applicable tools for the public health system of the region.

## Data Availability

Epidemiological data was provided for Directorate for Health Promotion and Prevention, Ministry of Health of the Santa Fe province, and the Epidemiology Division of the Entre Rios province.

## Acknowledgement

This research did not receive any specific grant from funding agencies in the public, commercial, or not-for-profit sectors. The authors would like to thank Directorate for Health Promotion and Prevention, Ministry of Health of the Santa Fe province, and the Epidemiology Division of the Entre Ríos province, for provided the epidemiological data. Also, the authors would like to thank National Meteorological Service (SMN) of Argentina, National Water Institute (INA)) and Argentine Naval Prefecture (PNA) for provided the hydrometeorological data.

## Author Contributions

Conceived of the research: A.G., M.S.L.

Model coding and programming support: L.L., A.G.

Processed data from remote sensing: W.S., M.S.L.

Extracted and processed data from the model, results discussion: A.G.; L.L., L.G.

Wrote the manuscript: A.G., M.S.L.

Reviewed the manuscript: G.M., W.S., L.L., L.G.

